# CORRELATION BETWEEN AGE-RELATED CHANGES IN SKIN MORPHOLOGY AND TROPHIC FUNCTION OF THE TRIGEMINAL NERVE

**DOI:** 10.1101/2022.11.13.22282259

**Authors:** A. K. Kolomiytsev, A. V. Bazilevich, M.V. Peshkov

**Affiliations:** Rostov State Medical University, Department of Pathology; Taganrog Department of Pathology

**Keywords:** aging, skin, trigeminal nerve, neuronal loss

## Abstract

Huge advancements in the field of cosmetology and numerous methods of skin restoration have kept the studies of skin aging continually relevant and up-to-date. Age-related changes in the physiological parameters of skin cover might be related to a trophic innervation disorder, which, in its turn, is caused by aging manifesting itself in the corresponding structures of the vegetative nervous system. The present article aims at describing this particular skin aging mechanism. Reduction of the amount of cells in *n. sensorius superior* of the trigeminal nerve has been proven to be connected with a decrease in the amount of cells in the innervated areas of skin. This process may be considered one of the mechanisms of aging realized as a systemic phenomenon.

## 1. Introduction

Studying of skin aging mechanisms continues to be relevant. Growing interest in this problem may be explained by the intensive development of cosmetology and various methods of restoring the physiological parameters of the skin cover.

At present, there are two types of skin aging: internal (chronological) and external (photoaging) [1, 2]. Each type has its own morphological features. It is believed that unlike chronological aging, which is a genetically determined process, photoaging directly depends on the degree of exposure to the UV irradiation and a genetically predetermined degree of pigmentation [3].

Despite various reasons, both types of aging are based on the same mechanisms related to a disorder of metabolism of collagen, the main structural component of the skin. Collagen production in the skin of elderly people compared to the young is decreased by an average 75%, but the level of collagen degradation (as in photoaging) is increased by 75%. There is a simultaneous decrease in the amount of collagen I and collagen III, and a decrease in the ratio of the amount of collagen III to collagen I, that correlates with a person’s age.

It was found, that the basis of skin aging process lies in the fundamental changes related to fibroblasts - the main cell population of dermis: their amount and functional activity [4, 5].

At the same time, the role of changes in the vegetative nervous system in the aging process of the body as a whole is just beginning to be studied [6]. The relationship between the age-related changes of the skin and the nervous system structures innervating it has not been investigated, although the trophic function of the nervous system has long been a matter of common knowledge [7]. The morphological changes in the trigeminal nerve nuclei depending on age have already been studied [8].

This article attempts to identify one of the possible skin aging mechanisms related to a disorder of trophic innervation, caused by enduring age-related changes in the corresponding structures of the vegetative nervous system.

## 2. Research purpose

The purpose of the research is to find out whether there is a relationship between age-dependent morphological changes in *n. sensorius superior* of the trigeminal nerve and the process of aging of skin areas innervated by the trigeminal nerve.

## 3. Materials and methods

The research was undertaken using autopsy material from people of various age groups who died from cardiovascular and cerebrovascular diseases. In each case, there were investigated:

1. The nuclei of the trigeminal nerve. Transverse fragments of the medulla oblongata of 3-4 mm thick at the level of localization of *n. sensorius superior* of trigeminal nerve were taken. Fragments of the tissue of the medulla oblongata were fixed in a 48% formalin solution for 48 hours, with the subsequent use of standard histological processing. Cross-sections of 5 μm thickness were made, painted with hematoxylin and eosin. There was the total number of 4 sections in each case. The number of neurons in the right and left nuclei of the trigeminal nerve was calculated at 420x magnification (the diameter of the field of view in this case was 0.32 mm), with the subsequent calculation of the mean value for each case.
2. The face skin areas in the zone of innervation of the middle branch of the trigeminal nerve, symmetrically right and left. The preparations were fixed in a 10% formalin solution, standard histological processing was made, and sections of 5 μm thickness were made. In each observation, 4 serial sections, painted with hematoxylin and eosin were made. The study of age-related changes in the skin was carried out by calculating the total amount of cells in derm in the field of view of the microscope at 420x magnification (diameter of the field of view, respectively, 0.32 mm).

There has been an additional study of immunohistochemical expression of Ki-67 in order to assess functional condition of skin cells and calculate the proliferation index. The proliferation level was estimated with the help of an index, which was calculated as a mean of the number of the labeled nuclei per 100 registered nuclei. Counting the labeled nuclei was carried out in the representative fields of view with a relatively even distribution of cells in the derm.

All observations were classified according to age groups.

## 4. Results

56 cases of death from cardiovascular and cerebrovascular diseases at the age from 32 to 98 years were studied. The cases were distributed according to the age groups as follows:

> 31-40 years -5 cases (male-2, female- 3),
>
> 41-50 years - 6 cases (male -3, female -3),
>
> 51-60 years - 8 cases (male - 5, female - 3),
>
> 61-70 years - 12 cases (male - 6, female - 6),
>
> 71-80 years - 11 cases (male - 6, female - 5),
>
> 81-90 years - 11 cases (male- 4, female- 7),
>
> 91-100 years -3 cases (male- 1, female- 2).
>
> When studying the nuclei of the trigeminal nerve, the following patterns were found:
>
> 1. The number of neurons in the right and left nuclei differs slightly; the difference is not statistically significant.
> 2. Minor fluctuations in the shape and size of transverse sections of the nuclei were found.
> 3. The signs of a progressive decrease in the number of neurons in *n. sensorius superior* of trigeminal nerve with age were found. In the age group of 31-40 years the mean number of neurons in the field of view was 14.2; in the age group of 41-50 years -13.67; in the age group of 51-60 years -12.5; in the age group of 61-70 years -9.42; in the age group of 71-80 years -9.1; in the age group of 81-90 years -8.2; in the age group of 91-100-years -9.3. The study revealed a 38.4% decrease in the number of neurons in *n. sensorius superior* of the trigeminal nerve in the age groups of 81-90 years and 91-100 years compared with age group of 31-40 years.
>
> Morphological skin changes were evaluated by the presence of dystrophic and atrophic changes. With growing age it was noted that the dystrophic changes were increasing. It was found, that in the age group of 31–40 years the mean amount of cells in skin sections in the microscope field of view was 48.8, in the age group of 41–50 years - 40.5, in the age group of 51–60 years - 40.12, in the age group of 61– 70 years -32.84, in the age group of 71-80 years -32.3, in the age group of 81-90 years -32.1, in the age group of 91-100 years -33.
>
> There were signs of a 33.6% decrease in mean amount of cell elements in the age groups of 81-90 years and 91-100 years compared with age group of 31-40 years.

Studying proliferation index of dermal cells with the help of assessment of Ki-67 expression has shown that fluctuations in the proliferative activity have no direct dependence on the age and, thus, are not statistically important.

## 5. Statistical studies

To evaluate the correlative relationships between the age of the dead people, the number of neurons in the nuclei of the trigeminal nerve and the amount of cells in the skin areas innervated by this nerve, a correlation and regression analysis with Pearson coefficient was performed. The following results were obtained:

1. For the relationship between the number of neurons in the nuclei of the trigeminal nerve and age, the correlation coefficient (r) is -0.519. The relationship between the investigated featured is inverse; the closeness (strength) of the connection on the Cheddock scale is noticeable.
2. For the relationship between the decreases in the amount of cells in dermis, the correlation coefficient (r) is -0.550. The relationship between the investigated features is inverse; the closeness (strength) of the connection on the Cheddock scale is noticeable.
3. For the relationship between the decrease in the number of neurons and the amount of cells in dermis, the correlation coefficient (r) is 0.697. The relationship between the investigated features is direct; the strength of the connection on the Cheddock scale is noticeable.

Thus, the obtained results should be considered statistically significant.

Due to the fact that a decrease in the number of neurons with age is not observed in all the investigated cases, as well as a decrease in the total amount of cell elements in the innervated areas of the skin, the Pearson’s X^2^ criterion, allowing evaluating the qualitative characteristics of the phenomenon under study, was additionally calculated.

The following results were obtained:

1. For the relationship between the number of neurons in the nuclei of the trigeminal nerve and age, the Pearson’s X^2^ value is 5.931 with a critical level for this sample of 5.34.
2. For the relationship between the decrease in the amount of cells in dermis and the age of dead, the criterion value is 7.098 with a critical level for this sample of 5.34.
3. For the relationship between a decrease in the number of neurons and a decrease in the amount of dermal cells, the criterion value is 18.103 with a minimum level of 4.22.

Thus, the results should be considered statistically significant, and it is obvious that the correlative relationship between changes in the nuclei of the trigeminal nerve and the development of atrophic processes in the innervated skin areas is much more expressed when compared with age correlations.

## 6. Conclusions

There was found a decrease in the number of neurons in the nuclei of the trigeminal nerve with aging. It was also confirmed that with aging the atrophic processes in skin, characterized by the decrease in the amount of cell elements, were observed. The revealed correlative relationship between the decrease in the amount of cells in n. *sensorius superior* of the trigeminal nerve and the decrease in the amount of cells in the innervated areas of the skin are statistically more expressed compared with age-related correlations, and therefore it may be considered to be one of the mechanisms for the realization of aging as a systemic process.

## Data Availability

All data produced in the present work are contained in the manuscript

## References

1. Yaar M., Gilchrest B.A. Skin aging: postulated mechanisms and consequent changes in structure and function // Clin. Geriatr. Med. 2001. Vol.17, No4. P.617–630.

2. Scharffetter-Kochanek K., Brenneisen P., Wenk J., Herrmann G., Ma W., Kuhr L., Meewes C., Wlaschek M. Photoaging of the skin from phenotype to mechanisms // Exp. Gerontol. 2000. Vol.35, No3. P.307–316. 29. Yaar M., Gilchrest B.A. Skin aging: postulated

3. Zorina A.I., Deyev R.V., Zorin V.L., Cherkasov V.R. Fibroblast-mediated skin aging. Possibilities of therapeutic correction. Experimental and clinical dermatocosmetology. No.5, 2011 p.43–51

4. Yusova Zh.Yu. Involutional skin changes with taking into account its type of aging. Scientific journal. Section: medicine. Pharmacy. 2012 No.22 (141) Issue 20/2, p.97–100.

5. Baumann L., Weinkle S. Improving elasticity: the science of aging skin. Cosmet. Dermatol. 2007; 20:168–172.

6. Davis E.A. and Dailey M.J. A direct effect of the autonomic nervous system on somatic stem cell proliferation? American Journal of Physiology-Regulatory, Integrative and Comparative Physiology. Vol. 316, N 1. doi.org/10.1152/ajpregu.00266.2018.

7. Kolomiytsev A.K. Systemic Structural Hypothesis of Aging // Advanced Studies in Biology. 2016. N1. P.17–25.

8. Kolomiytsev A.K. Changes in the nuclei of vagus trigeminal nerves depending on age // Higher educational establishments news. North Caucasion region. Section: Natural sciences. 2013. № 3 (175). P. 101–104.

